# Validation of the short Mood and Feelings Questionnaire in young adulthood

**DOI:** 10.1101/2021.01.22.21250311

**Authors:** Olga Eyre, Rhys Bevan Jones, Sharifah Shameem Agha, Robyn E Wootton, Ajay K Thapar, Evie Stergiakouli, Kate Langley, Stephan Collishaw, Anita Thapar, Lucy Riglin

## Abstract

**Background:** Depression often onsets in adolescence and is associated with recurrence in adulthood. There is a need to identify and monitor depression symptoms across adolescence and into young adulthood. The short Mood and Feelings Questionnaire (sMFQ) is commonly used to measure depression symptoms in adolescence but has yet to be validated in young adulthood. This study aimed to (1) examine whether the sMFQ is a valid assessment of depression in young adults, and (2) identify cut-points that best capture a DSM-5 diagnosis of depression at age 25.

**Methods:** The sample included young people who took part in the Avon Longitudinal Study of Parents and Children (ALSPAC) at age 25 (n=4098). Receiver Operating Characteristic analyses were used to examine how well the self-rated sMFQ discriminates between cases and non-cases of DSM-5 Major Depressive Disorder (MDD) classified using the self-rated Development and Well Being Assessment. Sensitivity and specificity values were used to identify cut-points on the sMFQ.

**Results:** The sMFQ had high accuracy for discriminating MDD cases from non-cases at age 25. The commonly used cut-point in adolescence (≥12) performed well at this age, best balancing sensitivity and specificity. However, a lower cut-point (≥10) may be appropriate in some contexts, e.g. for screening, when sensitivity is favoured over specificity.

**Limitations:** ALSPAC is a longitudinal population cohort that suffers from non-random attrition.

**Conclusions:** The sMFQ is a valid measure of depression in young adults in the general population. It can be used to screen for and monitor depression across adolescence and early adulthood.

## Introduction

Depression commonly onsets in adolescence (Kessler et al., 2005). It is the most common mental health problem, a leading cause of global disability, and is associated with repeated episodes of illness and poor long term outcomes including suicide (Costello and Maughan, 2015; Dunn and Goodyer, 2006; Harris and Barraclough, 1998; Kessler et al., 2005; Thapar et al., 2012). Therefore, accurate identification and monitoring of depression symptoms across adolescence and into young adulthood is important (NICE, 2019).

For robust examination of symptom stability and change in longitudinal research and in clinical practice, the use of the same measure at each assessment is required (Goodman et al., 2007). However, measures of depression that are commonly used for research and clinical practice in childhood and adolescence (e.g. Mood and Feelings Questionnaire, MFQ (Angold and Costello, 1987)) are different to the ones utilised in adult life (e.g. Beck Depression Inventory, BDI (Beck et al, 1961)). Whilst there is a need for measures that are validated across childhood, adolescence and into adulthood, research suggests differences in aetiology (Rice, 2010) and treatment (Thapar et al., 2012) across these age groups. Developmental considerations are important (Thapar and Riglin, 2020), including in clinical practice during the transition from child and adolescent mental health services (CAMHS) to adult mental health services (AMHS). It therefore cannot be assumed that the same validated childhood/adolescent questionnaires are also appropriate in adulthood.

The Mood and Feelings Questionnaire (MFQ) was originally designed to assess depression symptoms in children and adolescents (Angold and Costello, 1987). This measure is commonly used in clinic and research settings. It has been validated against DSM-IV and ICD-10 defined depression diagnosis (Daviss et al., 2006; Wood et al., 1995) and is a recommended screening tool for depression in children and young people (NICE, 2019). Although it does not have prescribed cut-points for all circumstances, specific cut-point thresholds validated against depression diagnosis have been proposed for the self-rated and parent-rated versions of this measure (Daviss et al., 2006; Wood et al., 1995).

A shorter version, the short Mood and Feelings Questionnaire (sMFQ) (Angold et al., 1995), is highly correlated with the MFQ (Thabrew et al., 2018) and has also been validated in both clinical and non-clinical samples (Thabrew et al., 2018; Thapar and McGuffin, 1998; Turner et al., 2014). It provides a quicker alternative to the MFQ for clinicians and researchers. Whilst the sMFQ has been validated at age 18 (Turner et al., 2014), to date its validity as a measure of depression post-18 in young adulthood has not been established.

This study aims to (1) examine whether the short MFQ is a valid assessment of depression in young adults, and (2) identify optimal cut-points for research and clinical practice that capture a DSM-5 diagnosis of major depression at age 25 years.

## Methods

### Sample

The sample was taken from the Avon Longitudinal Study of Parents and Children (ALSPAC), a well-established prospective, longitudinal birth cohort study. Pregnant women resident in Avon, UK, with expected dates of delivery 1st April 1991 to 31st December 1992 were invited to take part in the study. The initial number of pregnancies enrolled was 14,541 (for these at least one questionnaire has been returned or a “Children in Focus” clinic had been attended by 19/07/99). Of these initial pregnancies, there was a total of 14,676 foetuses, resulting in 14,062 live births and 13,988 children who were alive at 1 year of age. When the oldest children were approximately 7 years of age, an attempt was made to bolster the initial sample with eligible cases who had failed to join the study originally. As a result, the total sample size for data collected after the age of seven was therefore 15,454 pregnancies, resulting in 15,589 foetuses. Of these 14,901 were alive at 1 year of age. Part of this data was collected using REDCap (https://projectredcap.org/resources/citations/). Ethical approval for the study was obtained from the ALSPAC Law and Ethics Committee and Local Research Ethics Committees. Informed consent for the use of data collected via questionnaires and clinics was obtained from participants following the recommendations of the ALSPAC Law and Ethics Committee at the time. Please note that the study website contains details of all the data that is available through a fully searchable data dictionary and variable search tool: http://www.bristol.ac.uk/alspac/researchers/our-data/. Further details of the study, measures and sample can be found elsewhere (Boyd et al., 2013; Fraser et al., 2013; Northstone et al., 2019). For this study, data collected at age 25 years were utilised (n=4098). Where families included multiple births, we only included the oldest sibling.

### Measures

#### Short Mood and Feelings Questionnaire (sMFQ)

The self-rated sMFQ (Angold et al., 1995) was completed by ALSPAC participants at age 25 years. This is a 13-item questionnaire derived from the 33-item MFQ, designed for assessment of depression symptoms in children and adolescents. The questionnaire asks about depression symptoms in the last two weeks. Each item is rated on a 3-point scale (0=not true, 1=sometimes true, 2=true), with a total score ranging from 0-26. Higher scale scores suggest greater depressive symptoms.

#### Development and Well Being Assessment (DAWBA)

The self-rated DAWBA (Goodman et al., 2000) was used to derive a DSM-5 diagnosis of Major Depressive Disorder (MDD) in the sample at age 25. The DAWBA is a structured diagnostic measure that now includes items based on DSM-5. The depression section of the DAWBA asks about depression symptoms over the previous 4 weeks. Diagnosis coding was derived by OE, RBJ (both psychiatrists) and LR (psychiatry researcher). Diagnosis required the presence of five (or more) symptoms of depression (symptom present if participant responded “yes” to having the symptom) including either depressed mood or loss of interest or pleasure, plus evidence of distress or impairment (Table 1).

**Table 1.** Generating DSM-5 MDD diagnosis using self-rated DAWBA.

| **Five (or more) of the following symptoms have been present during the same 2-week period in the past 4 weeks and represent a change from previous functioning; at least one of the symptoms is either (1) depressed mood or (2) loss of interest or pleasure.** | **N with symptom/ N responding to question (%)** |
| --- | --- |
| *Core symptoms* |  |
| 1. Depressed most of the day, nearly every day for 2 weeks or more (very sad, miserable, unhappy or tearful) | 329/4082 (8.1%) |
| 2. Loss of interest in everything, or almost everything, normally enjoyed most of the day, nearly every day for 2 weeks or more | 324/4084 (7.9%) |
| **Meet criteria for (1) depressed mood or (2) loss of interest or pleasure** | **420/4077 (10.3%)** |
| *Other symptoms in those with core symptoms, during the same period* |  |
| 3. Eating much more or less than usual, or weight gain or loss | 332/420 |
| 4. Found it hard to get to sleep, stay asleep or slept too much | 388/420 |
| 5. Agitated or restless for much of the time | 309/420 |
| 6. Loss of energy or seemed tired all the time | 401/420 |
| 7. Feelings of worthlessness or unnecessary guilt for much of the time | 366/420** |
| 8. Found it hard to concentrate or think things out | 358/419** |
| 9. Thought about death a lot, talked about harming or killing self or tried to harm or kill self | 260/419** |
| **Meet criteria for five (or more) symptoms including either depressed mood or loss of interest or pleasure\*** | **395/4077 (9.7%)** |
| The symptoms caused upset or distress, or impairment in getting along with the people closest to (e.g. family, partner), making and keeping friends, work or study, or leisure activities. | 378/394*** |
| **Meet criteria for five (or more) symptoms including either depressed mood or loss of interest or pleasure plus evidence of distress or impairment** | **378/4076 (9.3%)** |
| *Loss of interest only counted as an additional symptom to depressed mood if they occurred at the same time. **Missing data for total N=3 (1 symptom missing each): all had ≥5 symptoms despite incomplete data. ***N=1 with missing impairment data: excluded. |  |

### Analyses

Data were analysed using Stata version 14. Where <10% items (1 item) were missing on the sMFQ, mean imputation was used to generate the missing value. Total sMFQ scores were calculated and mean total sMFQ scores were compared between those with and without DAWBA diagnosis of MDD. The internal consistency of the sMFQ was established by examining Cronbach’s alpha (values of ≥0.90 are considered excellent, 0.85-0.90 good, 0.80-0.85 moderate and 0.75-0.80 fair; Ponterotto and Ruckdeschel, 2007).

Receiver Operating Characteristic (ROC) analyses were used to examine how well the sMFQ discriminates between cases and non-cases of MDD classified using the DAWBA at age 25. A ROC curve was plotted (sensitivity vs 1-specificity), and the area under the curve (AUC) estimated. The AUC ranges from 0.5 to 1.0, with values of 0.5-0.7 usually interpreted as low test accuracy, 0.7-0.9 as moderate test accuracy, and > 0.9 as high test accuracy (Henderson, 1993). Sensitivity, specificity, positive predictive value (PPV) and negative predictive value (NPV) were derived. Sensitivity and specificity values were used to identify possible cut-points on the sMFQ at age 25.

The selection of specific cut-points on questionnaire measures is based on the context and rationale for using that measure, balancing the need for sensitivity versus specificity. Firstly, we aimed to identify a cut-point providing a good trade-off between sensitivity and specificity using maximal Youden Index (sensitivity + specificity −1; Fluss et al., 2005). This has been proposed as appropriate for a one-stage screening approach for epidemiological studies or research questions (Löwe et al., 2004). Secondly, we aimed to identify a cut-point more suitable for clinical screening of depression, where a two-stage approach might be used (i.e. the completion of the questionnaire is followed up by further clinical assessment). In such cases, it has been proposed that sensitivity should be high and be favoured over specificity, whilst retaining a specificity of at least 75% (Löwe et al., 2004). Given recent findings that lower cut-points on the sMFQ may be appropriate in males compared to females (Jarbin et al., 2020), we conducted sensitivity analyses running ROC analyses by sex.

## Results

### Descriptives

Of the n=4098 ALSPAC participants who took part at age 25, n=4085 completed the self-report sMFQ and n=4076 completed the DAWBA, with n=4063 completing both measures (99%). Of these, 66.6% (n=2707) were female and the mean age was 25 years (range 24-27).

A total of 9.3% (n=378/4076) of the sample met DSM-5 criteria for MDD at age 25 (Table 1). Female sex was associated with an increased likelihood of meeting MDD criteria (11.0% in females, 5.9% in males: OR=1.98, 95% CI=1.53-2.56, p=2×10^−07^). The mean sMFQ score for the whole sample was 6.83 (SD=6.40, range 0-26) and this was higher in females (mean=7.52, SD= 6.71) than males (mean= 5.44, SD=5.47), (t=-9.90(4083), p<0.001). The mean sMFQ score was higher in those with DAWBA MDD diagnosis (mean=17.47, SD=5.75) than in those without (mean=5.74, SD= 5.38) (t=-40.12(4061), p<0.001).

### Internal consistency

The Cronbach’s alpha for the sMFQ at age 25 was 0.92, suggesting the internal consistency of this measure was excellent.

### Receiver Operating Characteristic (ROC) analyses and criterion validity

ROC analyses suggested that the sMFQ had high accuracy for discriminating MDD cases from non-cases at age 25: AUC=0.92 (95% CI = 0.90-0.93). The ROC curve is shown in Figure 1. Sensitivities, specificities, PPVs and NPVs for a range of possible sMFQ cut-points that showed sensitivity and specificity >60% are shown in Table 2. Cut-points of ≥11, ≥12 and ≥13 all showed high sensitivity and specificity of >80%. Increasing specificity/PPV and decreasing sensitivity/NPV was observed with increasing cut-point values.

**Table 2.** Sensitivities, specificities, PPV and NPV for cut-points on sMFQ, compared against reference of self-rated DAWBA MDD diagnosis.

| **Cut-point** | **Sensitivity** | **Specificity** | **PPV** | **NPV** |
| --- | --- | --- | --- | --- |
| ≥ 6 | 97.4% | 60.2% | 20.1% | 99.6% |
| ≥ 7 | 96.0% | 65.9% | 22.4% | 99.4% |
| ≥ 8 | 93.1% | 70.8% | 24.6% | 99.0% |
| ≥ 9 | 90.2% | 74.7% | 26.8% | 98.7% |
| **≥ 10** | **88.9%** | **79.0%** | **30.3%** | **98.6%** |
| ≥ 11 | 86.0% | 82.4% | 33.4% | 98.3% |
| **≥ 12** | **83.6%** | **85.5%** | **37.1%** | **98.1%** |
| ≥ 13 | 80.2% | 87.8% | 40.2% | 97.7% |
| ≥ 14 | 73.5% | 89.9% | 42.8% | 97.1% |
| ≥ 15 | 69.3% | 91.9% | 46.8% | 96.7% |
| ≥ 16 | 65.1% | 93.2% | 49.5% | 93.6% |

Suggested sMFQ cut points are highlighted in bold. A cut-point of ≥ 12 best balanced sensitivity/specificity. A cut-point of ≥ 10 was most appropriate when favouring sensitivity over specificity.

**Figure 1:**
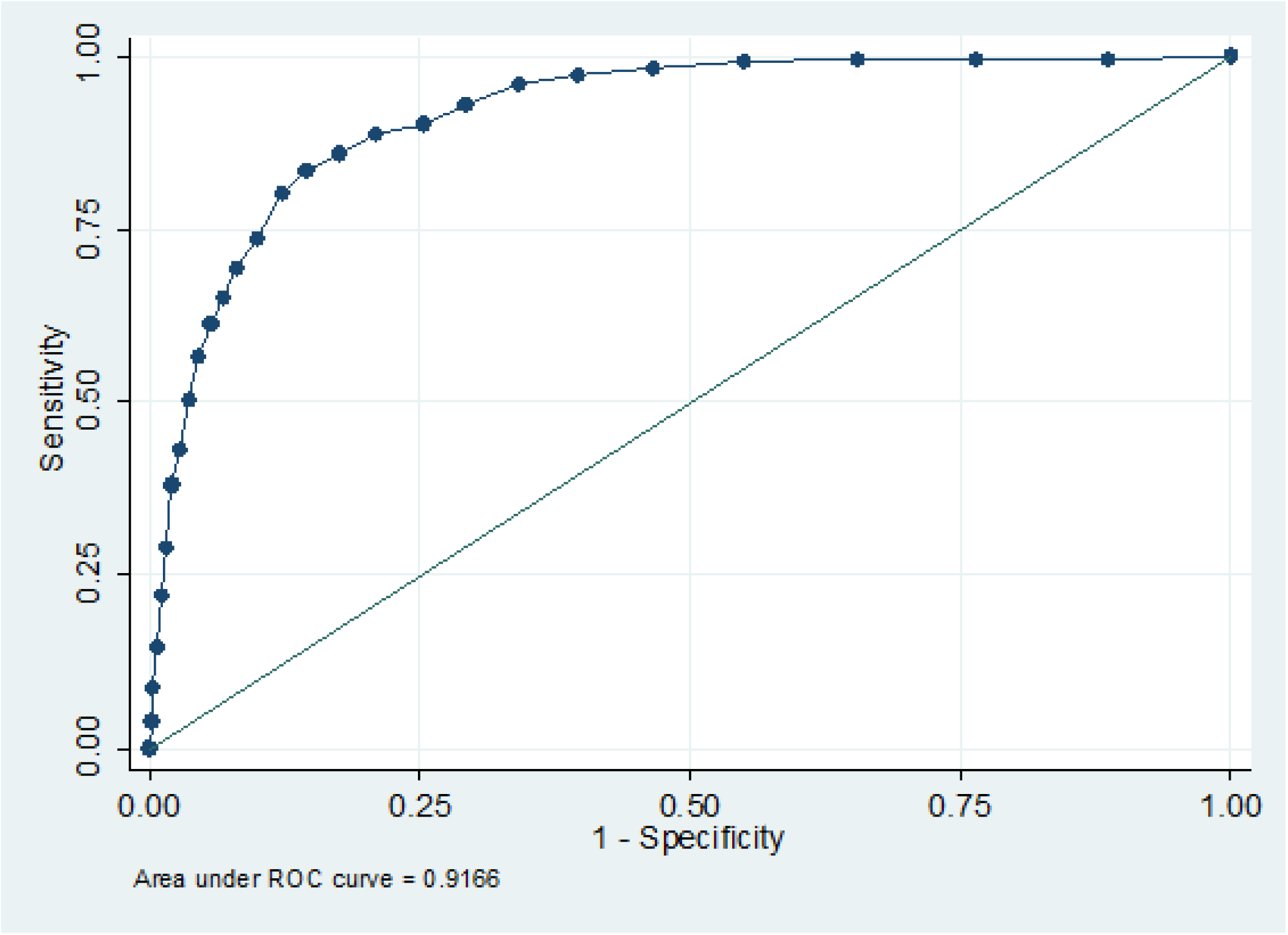
Receiver operating characteristic (ROC) curve for sMFQ using DAWBA MDD diagnosis as criterion.

When considering females and males separately, ROC analyses continued to show high accuracy, with AUC= 0.91 (95% CI = 0.90-0.93) for females and AUC=0.92 (95% CI = 0.89-0.95) for males and (see Figure 2). Table 3 shows the sensitivities, specificities, PPVs and NPVs for a range of cut-points for females separately.

**Table 3.** Sensitivities, specificities, PPV and NPV for cut-points on sMFQ, compared against reference of self-rated DAWBA MDD diagnosis by sex.

|  | Males |  |  |  | Females |  |  |  |
| --- | --- | --- | --- | --- | --- | --- | --- | --- |
| Cut-point | Sensitivity | Specificity | PPV | NPV | Sensitivity | Specificity | PPV | NPV |
| ≥ 6 | 95.0% | 67.2% | 15.4% | 99.5% | 98.0% | 56.5% | 21.8% | 99.6% |
| ≥ 7 | 92.5% | 72.1% | 17.2% | 99.4% | 97.0% | 62.6% | 24.3% | 99.4% |
| ≥ 8 | **91.3%** | **77.3%** | **20.1%** | **99.3%** | 93.6% | 67.3% | 26.2% | 98.8% |
| ≥ 9 | 86.3% | 80.9% | 22.0% | 98.9% | 91.3% | 71.5% | 28.4% | 98.5% |
| ≥ 10 | 86.3% | 84.9% | 26.3% | 98.99% | **89.6%** | **75.9%** | **31.5%** | **98.3%** |
| ≥ 11 | **83.8%** | **87.4%** | **29.4%** | **98.8%** | 86.6% | 79.8% | 34.6% | 98.0% |
| ≥ 12 | 78.8% | 90.2% | 33.5% | 98.5% | **84.9%** | **82.9%** | **38.1%** | **97.8%** |
| ≥ 13 | 73.8% | 91.8% | 36.0% | 98.2% | 81.9% | 85.6% | 41.4% | 97.4% |
| ≥ 14 | 65.0% | 93.3% | 37.7% | 97.7% | 75.8% | 88.2% | 44.2% | 96.7% |
| ≥ 15 | 58.8% | 95.0% | 42.3% | 97.3% | 72.2% | 90.3% | 47.9% | 96.3% |
| ≥ 16 | 55.0% | 95.9% | 45.8% | 97.1% | 67.8% | 67.8% | 50.4% | 95.8% |

Suggested sMFQ cut-points are highlighted in bold. For males, a cut-point of ≥11 best balanced sensitivity/specificity and a cut-point of ≥ 8 was most appropriate when favouring sensitivity over specificity. For females, a cut-point of ≥12 best balanced sensitivity/specificity and a cut-point of ≥10 was most appropriate when favouring sensitivity over specificity.

**Figure 2:**
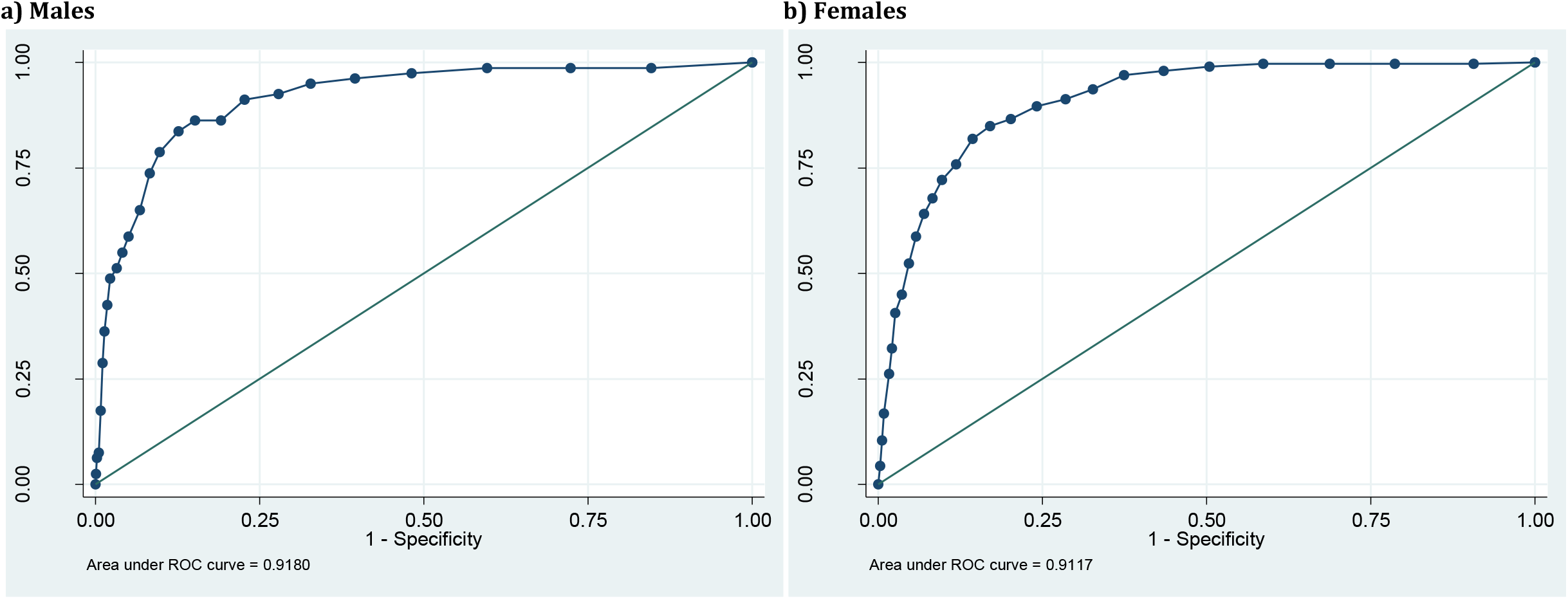
Receiver operating characteristic (ROC) curve for sMFQ using DAWBA MDD diagnosis as criterion, by sex.

### Optimal cut-points

Overall, a cut-point of ≥ 12 best balanced sensitivity/specificity on the sMFQ at age 25: sensitivity=83.6%, specificity=85.5%, PPV=37.1% and NPV=98.1%, according to the Youden Index (Fluss et al., 2005), capturing 21.2% (n=844) of the sample. When favouring sensitivity over specificity (as per Lowe et al, 2004), a cut-point of ≥ 10 was most appropriate: sensitivity=88.9%, specificity=79.0%, PPV=30.3%, NPV=98.6%, capturing 27.7% of the sample.

For males, an appropriate cut-point balancing sensitivity and specificity was ≥11 (sensitivity=83.8%, specificity=87.4%, PPV= 29.4% and NPV=98.8%), and for females ≥12 (sensitivity=84.9%, specificity=82.9%, PPV= 38.1% and NPV=97.8%). When favouring sensitivity over specificity an optimal cut-point for males was ≥8 (sensitivity=91.3%, specificity=77.3%, PPV=20.1% and NPV=99.3%) and for females was ≥10 (sensitivity=89.6%, specificity=75.9%, PPV=31.5% and NPV=98.3%).

## Discussion

This study aimed to use a population sample to examine the validity of the sMFQ in young adulthood, and to identify suitable cut-points for detecting a diagnosis of DSM-5 Major Depressive Disorder (MDD) in this age group. Our results show that the sMFQ is a valid measure of depression in young adulthood, with high accuracy in discriminating between cases and non-cases of MDD. Two possible cut-points were identified for screening for depression in different contexts.

The findings suggest that it would be acceptable to use the sMFQ when screening for depression in young adulthood. When compared to studies examining the validity of the sMFQ at earlier ages (Thabrew et al., 2018; Thapar and McGuffin, 1998; Turner et al., 2014), the sMFQ at age 25 performed very well at discriminating between cases and non-cases of MDD. The AUC in this study at age 25 years was 0.92 compared with AUC values of 0.67-0.87 for the self-reported sMFQ in childhood/adolescence (Rhew et al., 2010; Thapar and McGuffin, 1998; Turner et al., 2014).

The finding that the sMFQ is a valid screening tool for depression at age 25 also suggests it is a suitable measure for use across adolescence and into young adulthood, as it is already a widely used and validated measure of depression symptoms in adolescence (Thabrew et al., 2018). Our findings suggest it can also be used to measure stability and change in depression symptoms over time into young adulthood. This is useful for research settings, such as in longitudinal studies, but also in clinical settings where it may be useful to monitor depression symptoms across the transition from adolescence to adulthood (e.g. in primary care, or during transition from CAMHS to AMHS). The sMFQ also has the advantages that it only takes a few minutes to complete and is free to use.

We identified an sMFQ cut-point of ≥12 as optimal when balancing sensitivity and specificity. This cut-point is useful where a trade-off between false-positive and false-negative results is needed. For example, in research settings, where a decision on whether a young person may be depressed or not is made in one step. However, we also identified a second cut-point of ≥10, favouring sensitivity over specificity. This may be helpful, for example, when screening for depression in a clinical setting, where a questionnaire is followed by a further assessment (a two-step process) (Löwe et al., 2004). In these circumstances avoiding false negative results initially may be desirable.

Other studies of the self-reported sMFQ in younger samples have suggested widely ranging cut-points. Turner et al (2014) examined the validity of the sMFQ at age 18 years, also using the ALSPAC sample, but using a different depression diagnostic assessment tool (Clinical Interview Schedule-revised, CIS-R). They identified a similar cut-point of ≥ 11, although sensitivity was somewhat lower than we found (sensitivity 71.2%, specificity 83.0%, PPV 25.6%, NPV 97.2%). Rhew et al (2010) suggested a cut-point of ≥ 4 based on results from a school sample aged 11-13 years, Thapar et al (1998) suggested a cut-point of ≥ 8 in a twin sample aged 11-16 years, and Thabrew et al (2018) a cut-point of ≥ 12 in help-seeking 12-19 year-olds with mild to moderate depression. There is, therefore, some evidence that a lower cut-point may be appropriate in younger samples, with increasing thresholds needed for adolescents.

Variation in cut-points may also be related to the other sample characteristics, such as whether or not the young people are help-seeking and their sex. When we considered males and females separately, we found that lower cut-points on the sMFQ may be more suitable in males. This is supported by findings from studies in help-seeking adolescents. Jarbin et al (2020) suggested a lower cut-point for boys (≥6) compared to girls (≥12) and Thabrew et al (2018) suggested a cut-point of ≥12 for boys and ≥13 for girls. It is not clear to what extent age or puberty are responsible for the sex differences in cut-points.

### Strengths and limitations

This study has significant strengths. It is the first study, to our knowledge, to evaluate the validity of the sMFQ in adulthood, and utilises data from a large cohort of participants of a similar age.

However, there are limitations to consider. Firstly, ALSPAC is a longitudinal birth cohort that has been followed up over 25 years and suffers from non-random attrition, whereby some groups are more likely to drop out including those at elevated risk of depression as well as other psychopathology and lower socioeconomic status (SES) (Taylor et al., 2018). The sample is also a population sample, so generalising findings, especially the descriptive or demographic results, to other settings should be done with caution.

Another important consideration is the validity of the reference measure of depression diagnosis, against which the sMFQ is compared. In this study, DSM-5 MDD diagnosis was established using the DAWBA administered as a self-reported questionnaire rather than by clinical interview administered by trained interviewers, and this limited the information available to derive diagnoses. In addition, the DAWBA was originally developed as a child and adolescent measure, although it has been adapted for and used in adult life (https://www.dawba.info/; Findon et al., 2016), and diagnoses were derived using diagnostic criteria that are applicable to adults, including evidence of distress/impairment.

### Conclusion

We found the sMFQ to be a valid measure of depression in young adults in a general population sample, both for males and females. This suggests that the sMFQ can be used to screen for and monitor depression across adolescence and early adulthood in both research and clinical settings, enabling the monitoring of change and stability in symptoms over this transition period. However, it is still important to consider developmental differences even where the same measure can be used.

## Data Availability

The study website contains details of all the data that is available through a fully searchable data dictionary and variable search tool: http://www.bristol.ac.uk/alspac/researchers/our-data/.

## Acknowledgments

We are extremely grateful to all the families who took part in this study, the midwives for their help in recruiting them, and the whole ALSPAC team, which includes interviewers, computer and laboratory technicians, clerical workers, research scientists, volunteers, managers, receptionists and nurses.

